# The Monash Learning Health System Maturity Matrix: Codesign of a Tool to Measure and Guide Improvement in Complex Health System Behaviour

**DOI:** 10.1101/2025.04.09.25325486

**Authors:** Darren Rajit, Alison Johnson, Sandy Reeder, Dominique Cadilhac, Joanne Enticott, Helena Teede

**Author notes:** Joint senior author (Joanne Enticott, Helena Teede).

## Abstract

**Importance:** Learning Health Systems (LHS) have proven efficacy in catalysing healthcare improvement, but adoption and scale-up in complex healthcare systems remains challenging, with limited implementation guidance.

**Objective:** To measure alignment with LHS principles and guide LHS implementation, we aimed to codesign, refine and apply an LHS Maturity Matrix (LHS-MM) based on the Monash LHS framework.

**Design:** In this mixed methods study, our scoping review identified existing tools. We then applied the Double Diamond design and innovation model (discover, define, develop, deliver) in the development of the LHS-MM. Insights from engineering, prior tools, and the Monash LHS Framework were leveraged to adapt the LHS-MM. This was refined through codesign, and triangulation with evidence-based implementation frameworks. The LHS-MM was then delivered in a test case on stroke.

**Participants:** Codesign was conducted with subject matter experts (n=18), and end users of the LHS-MM (n=11).

**Setting:** Wbithin a high-income high quality national health system (Australia), across regional and urban settings.

**Outcomes:** A tool to measure implementation fidelity and alignment of healthcare system behaviours and processes with LHS principles, and guide organisations in effective LHS implementation for healthcare improvement.

**Results:** Tools uncovered in the discover and define phase emerged from the scoping review included the Cincinnati Network Maturity Grid. We adapted this tool to align to the Monash LHS framework. Codesign elevated the tool to focus on assessing complex systems behaviours aligned to LHS principles, with modification of assessment criteria, rating scales and scenarios for use. The LHS-MM assesses system-level behaviours across eight components on a numerical, five-point scale (1-5), visualised as a radar chart. Components include stakeholder engagement, priority identification, evidence-based information, evidence synthesis and guidelines, data systems, benchmarking, implementation, and healthcare improvement. Finally, in the deliver phase, a test case in stroke care revealed ratings from 4/5 (Established) to 5/ 5 (Transformative).

**Conclusion:** Through an iterative and evidence-informed codesign process, we have generated the Monash LHS-MM. Further research and government implementation is underway to operationalise the Monash LHS-MM to measure fidelity and guide LHS implementation, advancing the field both within and beyond the Australian healthcare system and globally. As an implementation guide and monitoring tool, it will be a pivotal ingredient inside implementation toolkits currently being developed worldwide, supporting LHSs to fulfil their promise and enable the next frontier of healthcare innovation.

## Introduction

Effective, sustained improvement in complex health systems has been elusive ^1,2^. Theory driven Implementation science frameworks such as the Consolidated Framework for Implementation Research (CFIR) ^3^ are often used as static determinant frameworks with limitations in operationalising delivery of improvement or catalysing iterative learning in complex systems over time ^4,5^. Learning Health Systems (LHS), ^6^ have emerged in this context as an evidence-based approach, aligned to the process domain of the CFIR, positing that complex systems change requires aligning people, data and culture ^7–10^. LHS approaches seek to close the evidence to practice gap, by moving beyond the ‘what’, to a focus on the ‘how’: that is how an improvement is contextualised, implemented, assessed, optimised and sustained for impact ^11^. Despite increasing interest, LHS research has been mainly theoretical, with relatively few high-quality empirical implementation studies, as captured in our recent scoping review ^12^. Additionally, LHS research often lacks codesign ^10^ with limited involvement of stakeholders or end-users in prioritising problems and designing solutions ^13^ and most are academically focused on technological^14^ or data^15^ aspects of the LHS, with limited investigation of complex system, or socio-technical factors ^12^.

The evidence-based Monash LHS framework underpinning this work was developed through codesign with health system stakeholders and community end-users (Figure 1) to address these limitations. This involved engaging with national and international experts, a systematic literature review analysing successful LHS case studies ^10^, followed by stakeholder engagement ^16^ and codesign ^9^. The Monash LHS framework recognised health systems as complex adaptive systems and positions iterative improvement as emergent system level *behaviours* with healthcare improvement arising from the accumulation of complex non-linear processes and interactions ^17^. It recognised these interactions across four evidence domains with eight LHS components (Table 1). Components are operationalised at the systems level through processes and tools that are integrated to elicit key emergent systems behaviours, aiming for iterative healthcare improvement (Table 1), referred here as the “LHS Cycle”.

**Figure 1:**
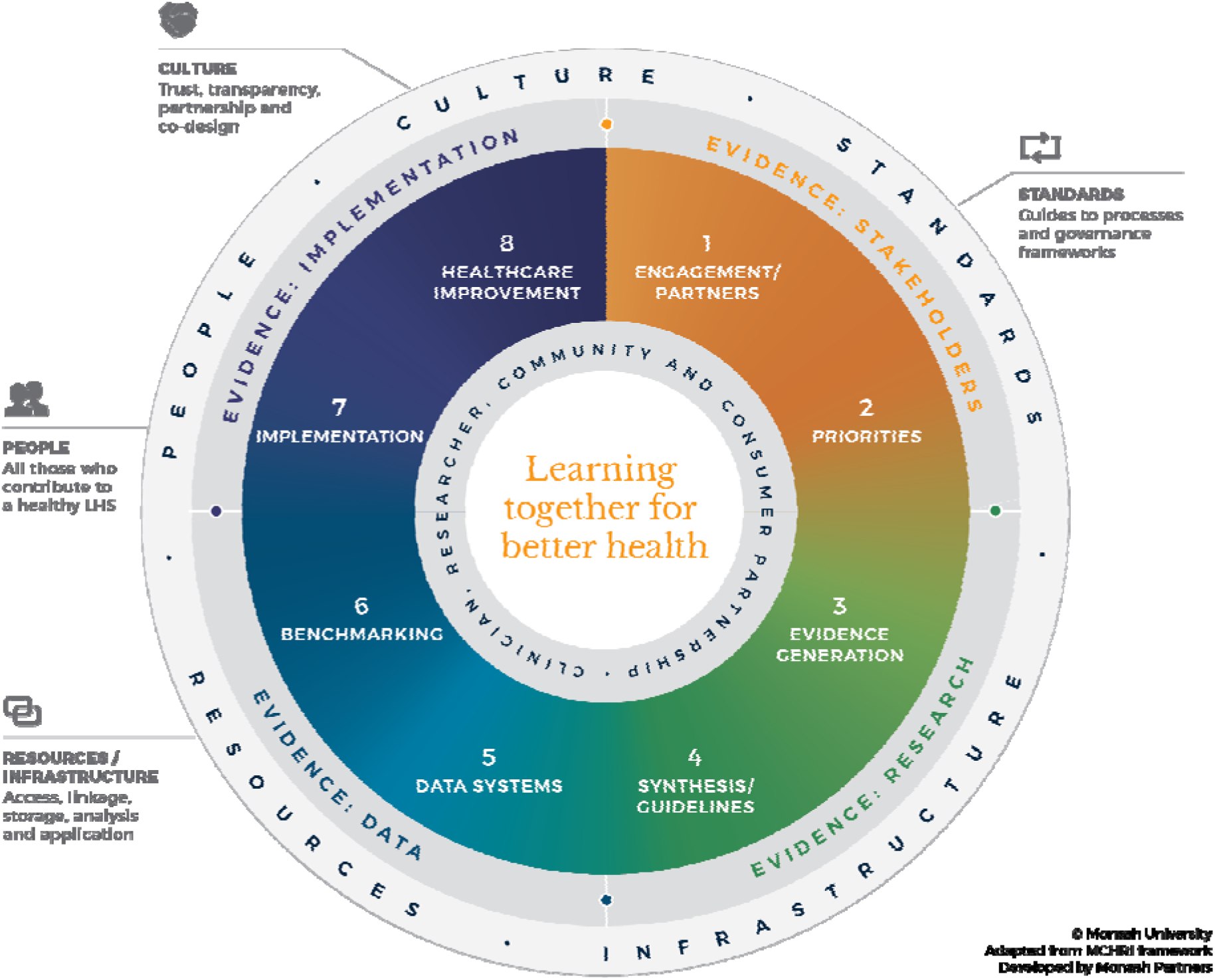
The evidence-based **Monash Learning Health System (LHS) Framework** ^9^. LHS Domains: Evidence from Stakeholders (orange), Evidence from Research (green), Evidence from Practice and Data (light blue), and Evidence from Implementation (dark blue). LHS Components are numbered.

**Table 1:**
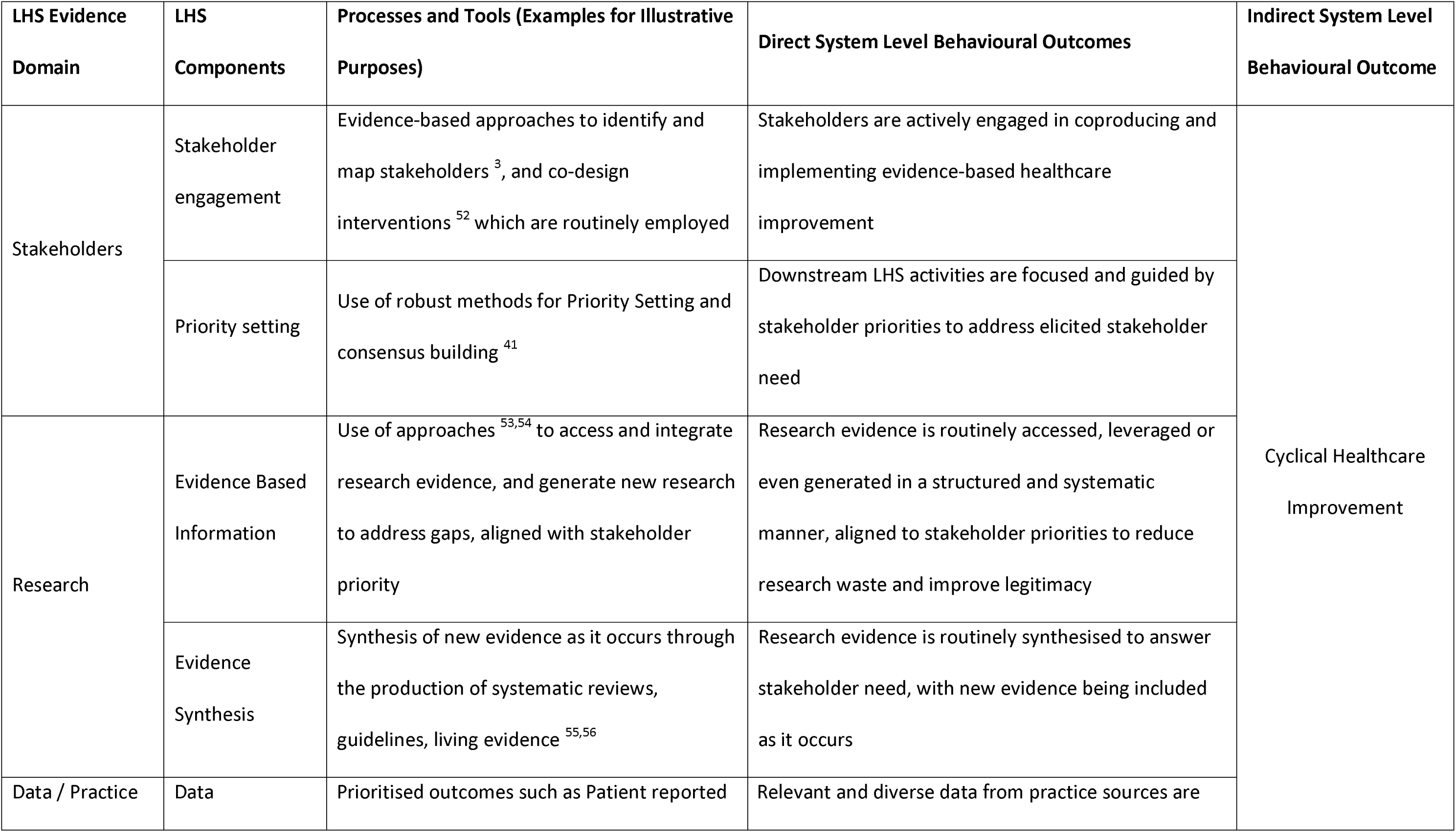
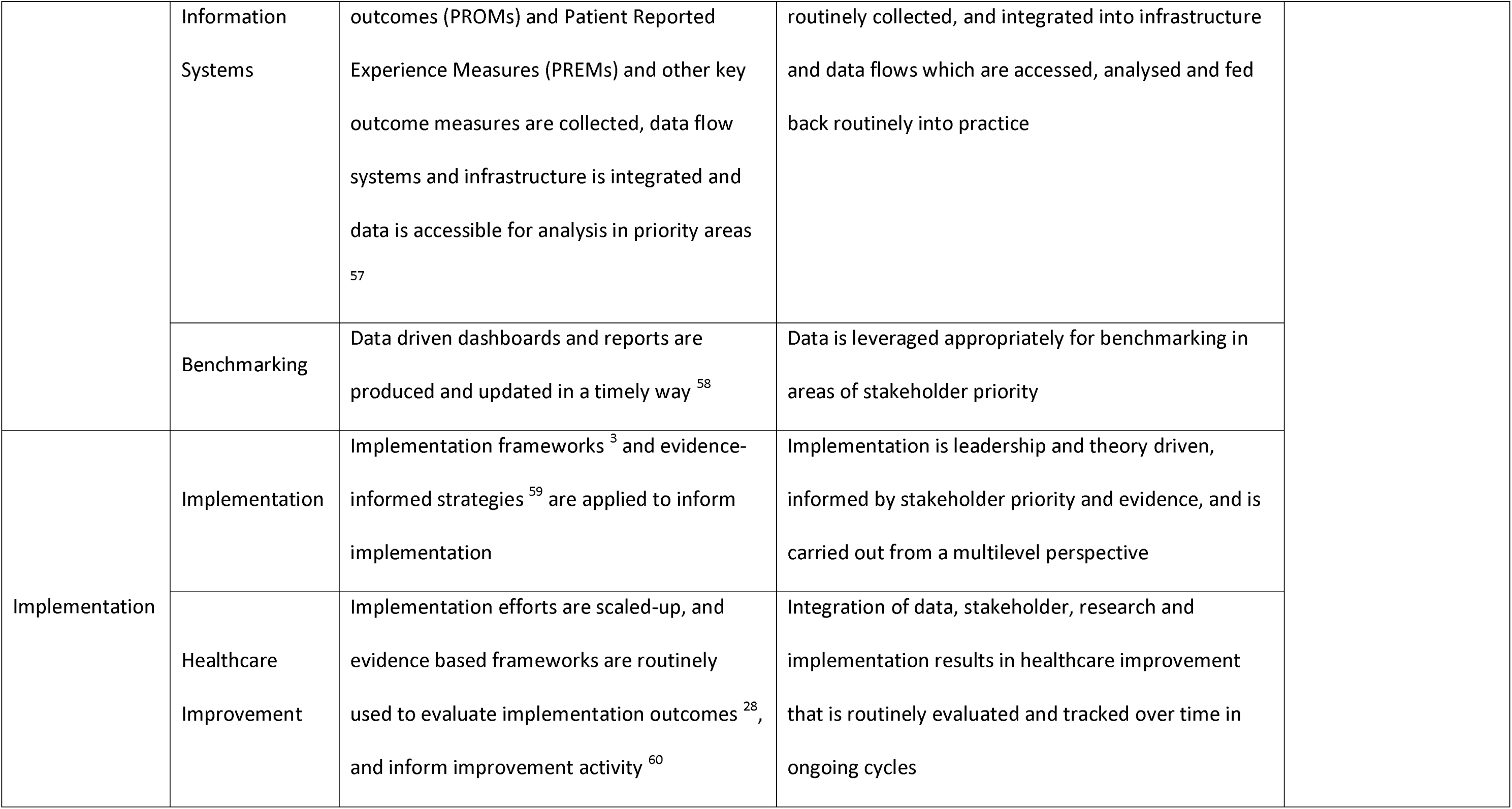
Examples of underlying processes or tools that are integrated and implemented in a Learning Health System to drive direct system level behavioural outcomes. Collectively, these direct behavioural outcomes lead to the higher level, indirect emergent behaviour of cyclical healthcare improvement.

The Monash LHS framework has been applied in Australia by government, health services and nationally funded implementation research and health system partnerships at urban (MRFF2023389 ^18^), regional (RARUR000072 ^19^) and national levels^7^. Ongoing LHS implementation and scale-up highlighted limited to monitor fidelity, benchmark or guide implementation. ^19^ We aimed to engage stakeholders to iteratively codesign an LHS Maturity Matrix (LHS-MM) within the Australian context, based on the Monash LHS framework.

## Method

This mixed methods study was underpinned by the validated codesign and innovation Double Diamond method ^20^, composed of four iteratively applied phases: Discover (Scoping Review, Expert and User Group Codesign), Define (Expert and User Group Codesign), Develop (triangulation with implementation science frameworks), and Deliver (Small-scale testing and refinement in the Australian Stroke LHS ^21,22^)

The work was led by our interdisciplinary team including a researcher-in-residence biomedical engineer (DR), clinicians (HT, DC, AJ) and implementation and LHS experts (HT, AJ, DC, SR, and JE). Ethics approval (ID:19969), Reporting and design of this study align with the Standards for Reporting Qualitative Research (SRQR) Checklist ^23^ (S6).

### Discover, & Define: Scoping Review, and Codesign Involving Experts and User Groups

An initial scoping review aiming to capture existing LHS implementation and evaluation research, including case studies and tools, was conducted and is published elsewhere^24^. The US based Network Maturity Grid tool ^25^ emerged as the most advanced^24^. It was developed over several years through evidence from stakeholders, a literature review, and multiple case examples ^25,26^. Here, we built on this tool in codesigning the LHS-MM, with permission for adaption provided from the creators.

A purposive sample of experts with experience in health systems research, LHS, evaluation, implementation science, health service delivery and complex research, as well as consumer and community members were engaged. The network established during the Monash LHS development (AJ, HT, JE) was also leveraged ^16^ The interview schedule was informed by our systematic reviews on effective LHS models ^10^ and scoping review on implementation tools ^24^ and was designed to discover the need, purpose, perceived usefulness, components and refinements of the tool, including exploring the skeleton prototype LHS-MM.

The interviewer (AJ) introduced the Monash LHS framework and context, before exploring the outlined topics and then the prototype LHS-MM. Input was solicited verbally and confirmed post interview via email and inline comments directly on prototype documents. The LHS-MM was revised iteratively, including around assessment criteria and maturity levels, evaluation matrix format, potential scenarios of use and target audiences, and possible integration into existing workflows. Successive dated versions of the tool stored on secure file servers. Interviews were continued to saturation, where no further actionable changes were added.

A workshop was conducted with identified stakeholders/ end-users, including embedded researchers and healthcare improvement project leads who were actively implementing the Monash LHS framework in a regional health system transformation program (MRFF RARUR000072) involving a diverse array of improvement projects. Participants were invited to apply the LHS-MM to assess LHS maturity in a group setting, with individual participants assessing each LHS component. Feedback and reflections were captured on experience using the LHS-MM, including potential usefulness in their broader work. Recommended refinements were captured via participatory engagement, with inputs captured verbally, in writing via email and directly on tool documents during and after the workshop.

All feedback from expert and end user engagement was considered of equal importance, aligned to sharing of power in co-design ^27^. Overall, multiple perspectives were captured and integrated to further define the tool. Throughout the codesign process, the co-authors met regularly and communicated via email to iteratively discuss and integrate evidence sources and build on the tool with successive dated versions stored on secure file servers.

### Develop: Triangulation with Theory Driven Frameworks

The LHS-MM generated via integrating results from the scoping review, feedback from experts and insights from the workshop, was then triangulated with evidence-based implementation frameworks (CFIR) ^3^, evaluation frameworks (Reach, Effectiveness, Adoption, Implementation, and Maintenance (RE-AIM)) ^28^, and the adherence construct of implementation fidelity ^29^. These frameworks were selected a-priori for their robust evidence base and extensive utilisation in implementation science^3,30–32^.

### Deliver: Small Scale Test Case – Australian Stroke LHS

The LHS-MM was then tested with the Australian Stroke LHS Program ^21^ based on publicly available evidence. This program ^21^ is a national network of consumer-clinician alliances ^22^, data monitoring systems ^33,34^, evidence synthesis bodies ^35^ and research centres ^36^ that have collectively led improvements in evidence-based stroke care in Australia. It was recently showcased as an exemplar for LHS approaches ^21^ aligning to the Monash LHS Framework ^9,22^. The test case was conducted independently by DR, with DC, a leader for the Australian Stroke LHS providing additional context, clarification, and evidence.

## Results

The multi-method results from each phase are summarised in Table 2. In summary, the scoping review generated a skeleton prototype that was adapted to the Monash LHS, with iteratively refinement during discovery, defining and developing the LHS-MM.

**Table 2:** Summarised results and contributions of each step in the multi method study to Monash LHS-MM development.

| Double Diamond Phase | Step | Contribution to LHS-MM Development |
| --- | --- | --- |
| Discover, Define | Scoping Review and adaptation of<br>the tool to Monash LHS framework | Uncovered the maturity grid assessment tool<br>(32) that formed the basis of further adaptation |
|  | Adapting an Implementation Fidelity Tool for the Monash LHS Framework | Basic structure of the Lannon et al. maturity matrix <sup>25</sup> was mapped to the Monash LHS Framework, resulting in an initial version for further iteration (S3) |
|  | Expert Feedback (n=18) | Changes in wording for maturity levels and assessment criteria, addition of options for different directions for usage, and interim version for focus group testing (S4) |
|  | User Group Workshop (n=11) | The workshop reaffirmed the LHS-MM, therefore the interim version (S4) proceeded to the next step |
| Develop | Triangulation with Theory Driven Frameworks | Insights from Process Domain of CFIR <sup>3</sup> and broad RE-AIM <sup>28</sup> constructs were integrated and led to wording changes to Implementation and Healthcare Improvement criteria, resulting in current version of Monash LHS-MM (S1-S2). |
| Deliver | Test case with the Australian Stroke LHS | Results from the Test case affirmed application of the LHS-MM as both an assessment tool and roadmap |

Collectively this codesign processes involved fundamental changes to structure, wording, rating criteria and conceptualisation of the LHS and its maturity (Summarised in S7).

### Monash LHS Maturity Matrix (LHS-MM)

The Monash LHS-MM tool includes a worksheet (S1) and report template (S2) and requires users to self-assess systems behaviours across four LHS domains and eight LHS components (Table 1), each with five maturity levels and associated quantitative scores: Not Started (1), Beginning (2), Developing (3), Established (4) and Transformative (5). These criteria (full wording in S1) evaluate system level behaviours generated through underlying processes that are characteristic of an increasingly mature LHS. At higher maturity levels, fulfilment of assessment criteria in a LHS domain is sequential and requires prerequisites from earlier domains. Complexity in assessment criteria increases with maturity, reflecting how LHS systems behaviours builds successively across components. This is especially apparent later in the Monash LHS cycle in the *Implementation* and *Healthcare Improvement* components. The result is a graphical view of LHS maturity (implementation fidelity) (Figure 2), highlighting strengths, deficiencies, and routes for improvement or investment.

**Figure 2:**
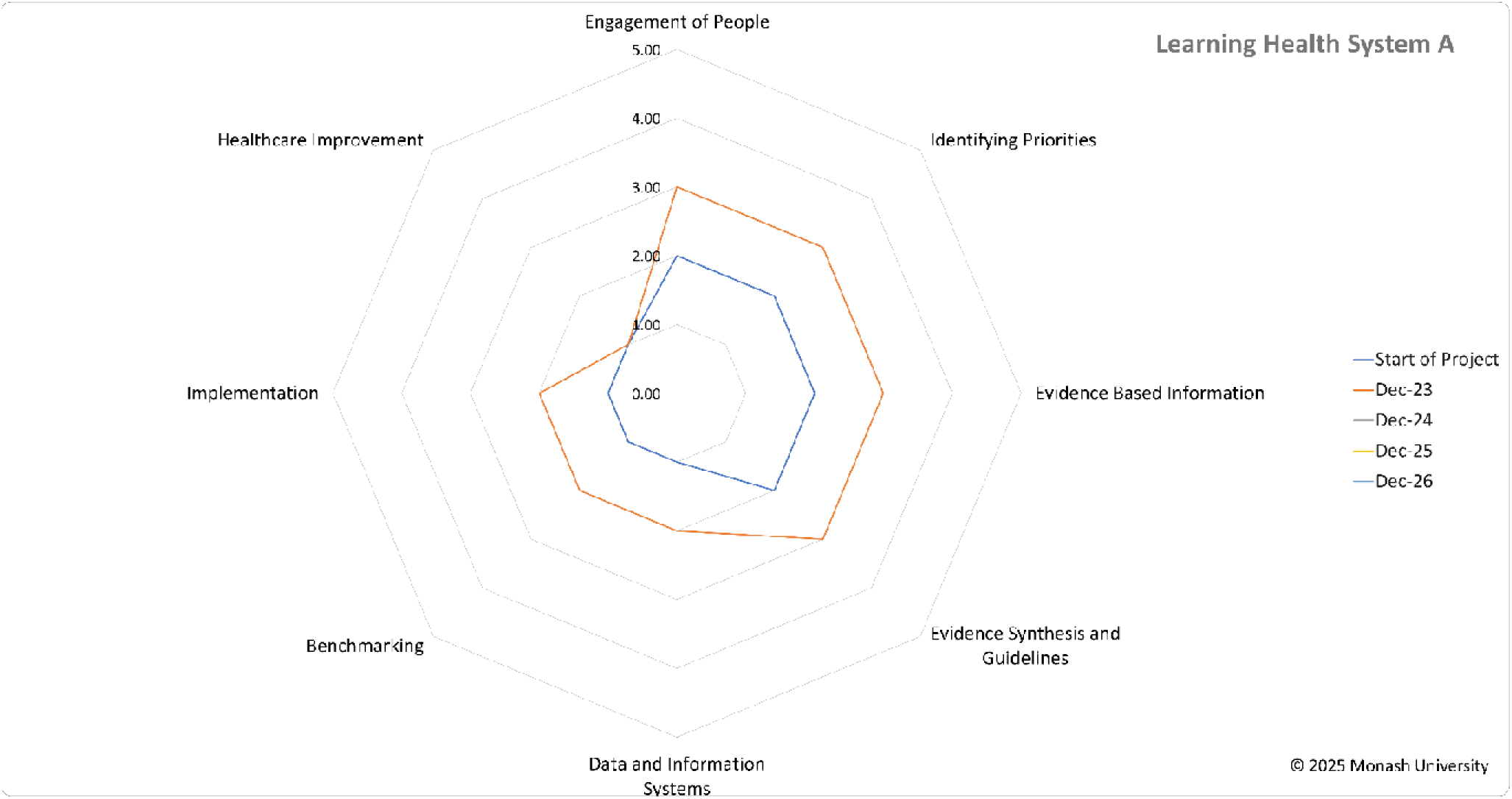
Sample Radar Chart that is produced by the Monash LHS Maturity Matrix (LHS-MM), highlighting ability to track maturity over time (Start of Project in Blue vs December 23 in Orange)

In the following sections, we provide further detail on assessment of LHS domains and components in the LHS-MM.

### Stakeholder Evidence Domain

Stakeholders derived evidence is generated from individuals or groups who have a stake in both the problems uncovered and the solutions that are proposed within a system ^37^. Measurement of maturity in this LHS domain focuses on how stakeholders are engaged *(LHS Component: Engagement of People*) for problem ideation, and co-design of interventions; and how stakeholder priorities are integrated into decision making and project planning *(LHS Component: Identifying Priorities)*. The *Engagement of People* scale is informed by the National Medical and Health Research Council framework for consumer involvement ^38^ and the International Association for Public Participation Spectrum of Public Participation model ^39^. The Identifying Priorities scale is informed by the James Lind Alliance approach to priority setting ^40^ and Delphi processes ^41^.

### Research Evidence Domain

Research derived evidence is generated by embedding high quality, peer-reviewed research. Measurement of maturity in this LHS domain focuses on how existing peer-reviewed research (*LHS Component: Evidence Based Information*) is being accessed, generated, incorporated and synthesised (*LHS Component: Evidence Synthesis and Guidelines*). At higher levels of maturity, integration of stakeholder priorities from the Stakeholder LHS domain is expected to guide evidence generation and synthesis, as well as new research evidence being integrated and synthesised as it is produced.

### Data and Practice Derived Evidence Domain

Data derived evidence refers to facts, circumstances or perceptions that can be analysed to inform decisions. This can be both qualitative and quantitative data that is generated through routine health system functioning. Measurement in this LHS domain focuses on how *relevant* data is being captured and accessed (*LHS Component: Data and Information Systems*); before being analysed, reported, and used to inform healthcare improvement activities (*LHS Component: Benchmarking*). Relevant data is determined by alignment with stakeholder priorities and research evidence, as elicited from the prior LHS domains. At higher maturity levels, there is clear evidence that stakeholder priority and research evidence is being used to drive the way data is being accessed, analysed and benchmarked to inform improvement activity in an ongoing cycle.

### Implementation Derived Evidence

Implementation derived evidence refers to i) how implementation science and practice-based evidence is being used to inform and create the conditions necessary for sustainable change and innovation (*LHS Component: implementation*), and ii) how evidence from the other three LHS domains are being integrated and evaluated (*LHS Component: Healthcare Improvement*) to underpin ongoing healthcare improvement. The *Implementation* component scale is informed by the CFIR framework, whereas the *Healthcare Improvement* component scale is informed by the RE-AIM framework. At higher maturity levels, implementation science and evidence from all LHS domains is being used at scale, to i) select and adapt novel innovations (Innovation Domain of CFIR), and ii) co-design and operationalise implementation or de-implementation strategies for these innovations (Process Domain of CFIR). Thereafter, evaluation from implementation activities is continuously used to inform cycles of healthcare improvement, allowing evidence from all LHS domains to be linked towards outcomes. This marks the end of one LHS cycle and the start of another.

### Monash LHS-MM as an Implementation Fidelity Assessment Tool

The LHS-MM is designed to assess the four subcategories (Content, Frequency, Duration and Coverage) of the “Adherence” construct of implementation fidelity in relation to the Monash LHS framework (Table 3). Implementation fidelity refers to the extent to which a complex intervention (the Monash LHS framework) has been implemented as intended by the developers, whereas “Adherence” is the “bottom line” of Implementation fidelity, *i.e.,* extent to which those wishing to implement LHS principles have “adhered” to the Monash LHS framework as planned. Adherence is decomposed into four subcategories: Content, Frequency, Duration, and Coverage. “Content” refers to “what” is being assessed, in this case how the Monash LHS framework has been conceptualised into measurable attributes as detailed above. Frequency, Duration and Coverage collectively refer to the “dose”, or the extent to which the Monash LHS framework is being delivered.

**Table 3:** The Monash LHS-MM approach to measure the Adherence construct of Implementation Fidelity, as related to the Monash LHS Framework.

| Adherence Subcategories | Monash LHS-MM Assessment Approach |
| --- | --- |
| Content | Breaks down the Monash LHS Framework and LHS cycle into system level behaviours across all LHS components and domains. Each component is assessed across five maturity levels, with criteria building on preceding maturity levels within LHS components, and between LHS components in other domains. |
| Frequency & Duration | Frequency is how often a LHS cycle is completed across all eight LHS components, and duration is how long each LHS cycle takes. Both of this can be measured via successive usages of the tool due to its flexible format. For example, an implementation fidelity assessment could be run every year, with each assessment over a one-month period. Sequential radar charts can then be overlaid to display evolution of adherence to the LHS model over time. |
| Coverage | “Coverage” is the extent, or depth to which the LHS has been delivered. Here the Monash LHS-MM measures this on a maturity scale from 1 – 5. |

### Test Case – The Australian Stroke Learning Health System Program

Figure 3 summaries the overall LHS maturity level of the Australian Stroke LHS. A completed LHS-MM, accompanying report, and assessment rationale with evidence for each component is available (S5a-SSc). Notably, the Australian Stroke LHS was rated as “Transformative (5/5)” in terms of Stakeholder engagement, Evidence Based Information, Evidence Synthesis, Data Information Systems, Benchmarking and Implementation. However, “Identifying Priorities” was an area for improvement (Developing (3/5)) due to limited evidence for how stakeholder priorities were being ranked. Additionally, given that implementation of tools and processes associated with the LHS principles was still relatively nascent, evidence of evaluation of ongoing LHS cycle is still nascent, thus a rating beyond Established (4/5) could not be assigned.

**Figure 3:**
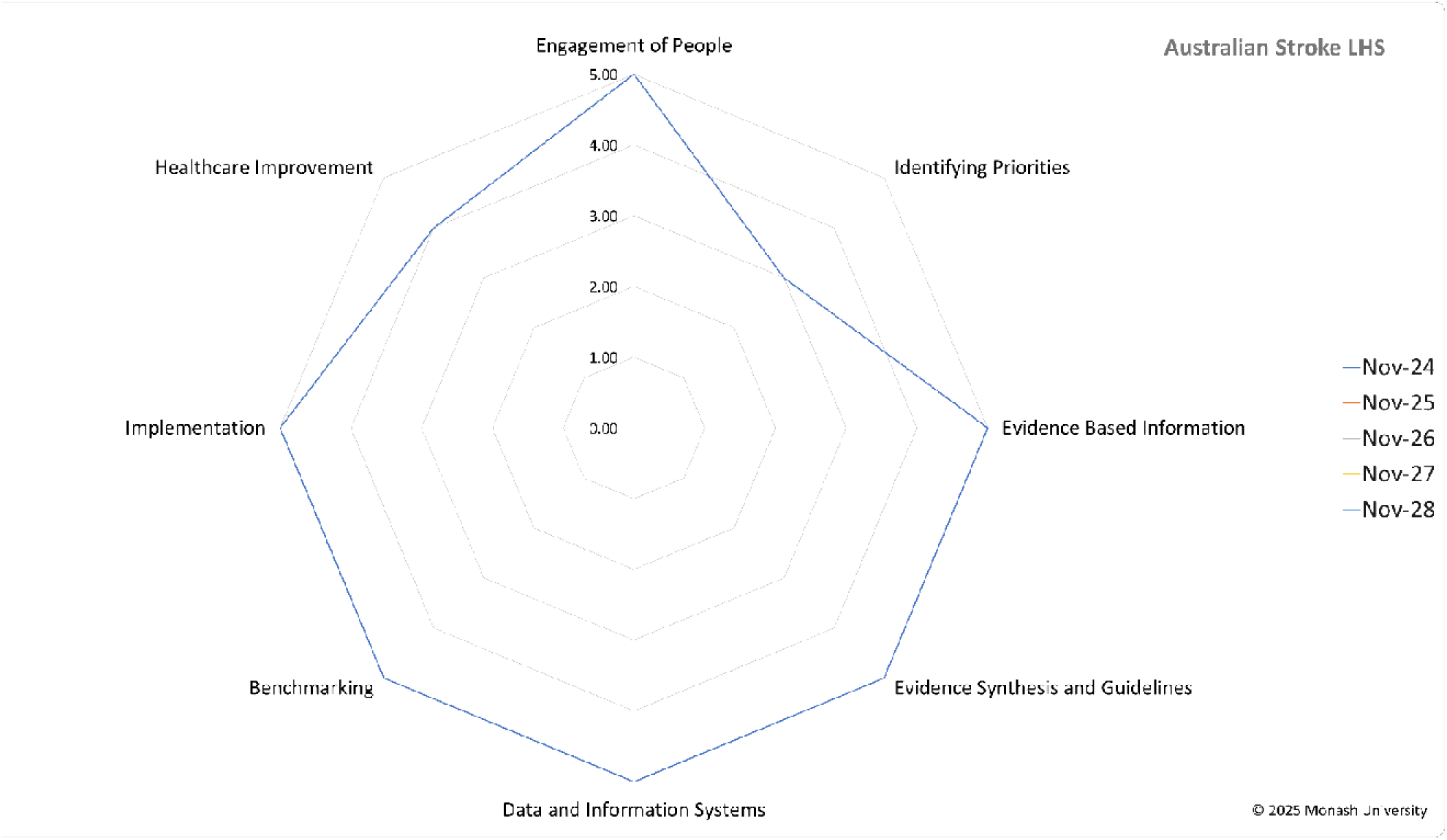
Radar chart summarising results of assessment of the Monash LHS Maturity Matrix (LHS-MM) of the Australian Stroke Program.

### Application

The LHS-MM can be used as a brief reflexive, analysis with documentation to support maturity scores as done in our test case (S5a-c), or a deep analysis using mixed methods data collection such as interviews; for example with a realist evaluation ^42^. Following the instructions in the LHS-MM worksheet (S1), the scores are completed and a summary sheet with a radar chart quantitatively visually displays maturity scores across all eight components (Figure 2) accompanied by a report template (S2) to record results. The tool can be applied using simple word and excel outputs and has also been incorporated into an online Monash LHS implementation toolkit, with further instructions and support information. The maturity assessment using the LHS-MM is best conducted iteratively with benchmarking over time and across organisations to guide targeted efforts into improving health systems behaviours towards a mature LHS.

## Discussion

Building on the LHS, we have generated a Monash LHS-MM codesigned from i) evidence from a scoping review of existing frameworks and tools ^24^ ii) codesign with expert stakeholders and end-users through the discovery, defining and development phases including integration of evidence-based, theory driven frameworks, (CFIR ^3^, RE-AIM ^28^ and the conceptual framework of Implementation Fidelity ^29^). The resultant LHS-MM, designed for measuring fidelity and guiding implementation of an evidence-based LHS framework to enhance healthcare improvement, was then delivered in a test case in the Australian LHS Stroke program ^21^. As such, it is both an implementation guide and monitoring tool, and can assist health services globally to establish a LHS to enhance healthcare improvements and deliver impact.

This codesign process for the LHS-MM was underpinned by the conceptualisation of the LHS as a series of measurable, system level behaviours. These behaviours are generalisable across contexts but delivered through flexible processes *unique* to context. To address the need to guide and measure LHS implementation, we build upon work originating in the United States on maturity matrices usage in LHSs (Network Maturity Grid) ^25^ ^43^. These have emerged from the engineering sector ^44,45^ to assess and optimise behavioural outcomes from a systems perspective (key divergences in S7), and have also been used in public health settings with similar large scale health transformation projects ^46–48^. Maturity matrices rely on behaviourally anchored rating scales ^49^ where each level on the scale depicts model behaviour to which assessors can compare their own context.

The LHS-MM includes quantitative rating scales to assess LHS system behaviours by maturity from 1 to 5, in all eight LHS components (Table 3). Here, we focus on assessing behaviours, without prescribing specific processes, allowing these to be employed in a bespoke manner, depending on context. This allows the flexible application of the LHS-MM across a range of settings and contexts in healthcare. Throughout the codesign process, the matrix was perceived to be useful in understanding, planning, implementing, refining and evaluating LHS implementation fidelity and adherence to the LHS framework. Next steps in development include implementation and evaluation in large scale national programs (APP1198561, APP2018718), which are underway across a range of LHS initiatives, with embedded research and evaluation ^50^.

The Monash LHS-MM builds on contemporary maturity matrices ^25,46,48,51^ and the LHS to integrate evidence-based implementation frameworks (CFIR^3^ and RE-AIM^28^) to improve system level capability in integrating implementation science at scale. For example, where CFIR excels at helping implementors identify “what” contextual, multi-level factors are salient to implementation, the LHS-MM helps implementors to quantitatively assess and chart a course to developing system level capabilities and behaviours that enable successful implementation and improvement activity; in this case both the “how” and “how well”. For example, in our Australian stroke test case, a lack of maturity in engaging stakeholders and enhancing priority setting was identified, and future investment could be targeted towards improving how stakeholder priorities are elicited via consensus, ranked, and shortlisted to inform downstream research, data and benchmarking, and implementation activity.

The major strength of the Monash LHS-MM is that it is a codesigned resource and can function as an implementation guide. As both implementation guide and monitoring tool, it will assist health services globally to establish a LHS to enhance healthcare improvements and deliver impact. It will be an important component in implementation toolkits currently being developed around the world to enable the implementation of learning health systems to continuously learn from practice, adapt interventions, and optimize outcomes. This will ensure that LHSs are move beyond conceptual models into practical, scalable systems that drive meaningful change. As part of such implementation toolkits, it will help bridge the gap between research and practice, allowing health services worldwide to achieve sustained improvements in patient care, operational efficiency, and health equity. Ultimately, the LHS-MM will contribute to transforming healthcare systems into dynamic, learning organizations capable of meeting evolving challenges and improving population health outcomes.

There are several limitations to this study. The LHS-MM was codesigned to align with the Monash LHS Framework and the Australian context. Therefore, it may require adaptation for other LHS models and settings. In refining the LHS-MM it was delivered through an Excel worksheet and Word template, which may lack sophistication, but enhances accessibility. It is now being integrated in an online implementation toolkit, with codesigned embedded guidance and resources. Workshops with end-users were conducted in group settings. As such, in-group dynamics may have influenced the results. Further, there is an element of subjectivity to self-assessment. Whilst the LHS-MM aims to enable a reflexive and self-assessment approach, this does allow potential bias, with the need for more prescriptive description of maturity that evolved over the codesign process. Further in-depth case studies are needed using mixed methods and incorporating multiple perspectives across a range of settings, to assess interrater reliability and construct validity. This is underway across multiple large-scale complex funded initiatives underpinned by the CFIR, LHS and the Monash LHS-MM.

## Conclusion

Learning Health Systems (LHS) aim to deliver healthcare improvement through eliciting and integrating evidence-based systems behaviours that are operationalised by a series of processes unique to context. Given their complexity, implementation can be challenging and tools to support LHS implementation and evaluate implementation fidelity are scarce. In response, we have codesigned the Monash LHS-MM through a scoping review, and a four phase Double Diamond codesign process involving stakeholder codesign, integration of evidence-based frameworks, and a test case. The resultant LHS-MM provides a structured approach to assess LHS implementation fidelity and maturity over time, supporting stakeholders to assess and optimise approaches to healthcare improvement. The LHS-MM has now been integrated into an online platform aligned with the CFIR and is being deployed across initiatives to benchmark and drive large-scale healthcare systems change. As both an implementation guide and monitoring tool, the LHS-MM will be a pivotal ingredient inside implementation toolkits currently being developed worldwide, supporting LHSs to fulfil their promise and enable the next frontier of healthcare innovation.

## Supporting information

S1_MonashLHS-MM_Worksheet_Template

S2_MonashLHS-MM_Report_Template

S5a_MonashLHSMM_AustralianStrokeLHS_Filled

S5a_MonashLHSMM_AustralianStrokeLHS_Report_Filled

S6_SRQR_Checklist

S7_MonashLHSMM-NMG-Divergence

## Data Availability

All data supporting the findings of this study is available within the paper and the accompanying supplementary information.

## List of Abbreviations

LHS: Learning Health System
LHS-MM: Learning Health System Maturity Matrix
CFIR: Consolidated Framework for Implementation Research
RE-AIM: Reach, Efficacy, Adoption, Implementation and Maintenance
BARS: Behaviourally Anchored Scales
SRQR: Standards for Reporting Qualitative Research
ASHC: Academic Health Science Centres
PROMs / PREMs: Patient Reported Outcome/ Experience Measures
PBS: Pharmaceutical Benefits Scheme
MBS: Medicare Benefits Schedule
ASC: Australian Stroke Coalition
AuSDaT: Australian Stroke Data Tool
AuSCR: Australian Stroke Clinical Registry (AuSCR)
DR: Darren Rajit
AJ: Alison Johnson
DC: Dominique Caldilhac
SR: Sandy Reeder
JE: Joanne Enticott
HT: Helena Teede

## Declarations

### Ethics approval and consent to participate

All interviews were recorded, and verbal consent provided at the beginning of the interview. This study was approved by the Monash University Human Research Ethics Committee (Project ID: 19969) who approved the method for verbal consent which is explained in the methods section.

Potential participants were invited to take part in the study by an introductory email, and then followed-up by a project researcher to provide study information (including prototypes of the matrix), answer questions and organise mutually agreeable interview times. Verbal consent to recording was obtained before commencing the interview.

### Consent for publication

The authors declare their consent for publication.

### Author Contributions

D.R, A.J, J.E and H.T contributed to conceptualisation. H.T obtained funding. A.J conducted the interviews. J.E and H.T contributed to supervision. D.C and S.R provided feedback on the maturity matrix during its development stage and D.C corroborated the stroke test case details. D.R. drafted the manuscript with H.T, and all authors contributed intellectually, revised and approved the manuscript.

## Acknowledgements

The authors would like to acknowledge the input and valuable feedback contributed by the subject matter experts and potential users of the tool that were engaged as part of maturity matrix development. Further, this work has leveraged the Deliver initiative (MRFF RARUR000072) aiming to leave a sustainable learning health system and resultant legacy of better health outcomes and a greater research capacity in regional settings in Australia.

