## Supplementary material for "The Monash Learning Health System Maturity Matrix: Codesign of a Tool to Measure and Guide Improvement in Complex Health System Behaviour": S2_MonashLHS-MM_Report_Template

### Learning Health System Matrix Snapshot report

This document will provide a system/ structure for documentation of use of the matrix over time.

#### Title of project/ service or organisation

**Date of evaluation**:

**Person completing the evaluation**:

**Role**:

#### Step 1:

Complete your matrix rating. (Worksheet 1. Make sure that you alter the cells on row 2, column H, I, J, K, L. This will ensure that the Legend on the spider graph is relevant to your study.)

Then copy and paste your spider graph (From the Summary Sheet) **to replace the one below**.

Learning Health System Maturity Matrix

|  | Maturity score - Project Start | Maturity score - March 2023 | Maturity score - March 2024 | Maturity score - March 2025 | Maturity score - March 2026 |
| --- | --- | --- | --- | --- | --- |
| Engagement of People | 2 | 3 | 0 | 0 | 0 |
| Indentifying Priorities | 1 | 2 | 0 | 0 | 0 |
| Evidence Based Information | 2 | 3 | 0 | 0 | 0 |
| Evidence Synthesis and Guidelines | 2 | 3 | 0 | 0 | 0 |
| Data and Information Systems | 1 | 2 | 0 | 0 | 0 |
| Benchmarking | 1 | 1 | 0 | 0 | 0 |
| Implementation | 1 | 2 | 0 | 0 | 0 |
| Healthcare Improvement | 1 | 2 | 0 | 0 | 0 |

#### Step 2:

Now record which approach you used to evaluate each of the eight components.

Place an x in the appropriate cell to indicate which approach you took (delete and re-enter the data in the table below as appropriate)

- Instinctual - This is a quick rating. Don't worry too much. Just fill in without reference to anything. Later steps will use a more evidence-based approach. This rating method will familiarise you with the process
- Evidence-based with reference to local information/ data - In this approach you use data and information that you have available. This may be surveys, monthly reports, emails. It can be any information that you can refer to that is relevant to your site/ project/ service
- Evidence based with use of evidence-based tools (those provided in the matrix and any others you have used instead) - The tools/ links that are provided are those recommended by our Stakeholders. Choose an area and then refer to the tools. Choose one to start with and see if your approach is consistent with the provided evidence-based approach. Overtime you would plan to look at each of the 8 areas with this approach.

Then indicate how many components were listed in each column.

Add in any notes and or recommendations here.

| **Evidence base** | | | | |
| --- | --- | --- | --- | --- |
| **Domain** | **Component** | **Reflexive** | **Evidence based with reference to local information/ data** | **Evidence based with use of evidence-based tools** |
| **Stakeholder Derived Evidence** | **Engagement of People** | x |  |  |
|  | **Identifying Priorities** | x |  |  |
| **Research Derived Evidence** | **Evidence Based Information** | x |  |  |
|  | **Evidence Synthesis and Guidelines** | x |  |  |
| **Data Derived Evidence** | **Data and Information Systems** | x |  |  |
|  | **Benchmarking** | x |  |  |
| **Implementation Evidence** | **Implementation** | x |  |  |
|  | **Healthcare Improvement** | x |  |  |
| **Count of evidence approaches** |  | 8 | 0 | 0 |

##### Step 3:

##### Evidence based with reference to local information/ data or other evidence-based tools

Where you have accessed some sort of evidence to complete your ranking it is advised to document clearly what they were. Over time it is easy for this information to be lost.

- Document which data sources, tools and resources you used in each of the component areas
- Indicate whether they were sourced from the spreadsheet or whether it was found elsewhere
- Include links to the source - or
- Attach any PDF’s if the data source is likely to change over time
- List information about the data/ information next to the relevant component as listed below:

###### Engagement of People

###### Identifying Priorities

###### Evidence Based Information

###### Evidence Synthesis and Guidelines

###### Data and Information Systems

###### Benchmarking

###### Implementation

###### Healthcare Improvement

#### Step 4:

On the following page is a blank repeat of the above. Remember to copy and paste in the blank one again, prior to completing at your next time point.

#### THIS IS YOUR BLANK TEMPLATE FOR FUTURE USE – COPY AND PASTE THIS TO THE START OF THE DOCUMENT WHEN COMPLETING YOUR NEXT REVIEW

#### Deliver

**Date of evaluation:**

**Person completing the evaluation**:

**Role:**

#### Step 1:

Complete your matrix rating. (Worksheet 1. Make sure that you alter the cells on row 2, column H, I, J, K, L. This will ensure that the Legend on the spider graph is relevant to your study.)

Then copy and paste your spider graph (From the Summary Sheet) **to replace the one below**.

Learning Health System Maturity Matrix

|  | Maturity score - Project Start | Maturity score - March 2023 | Maturity score - March 2024 | Maturity score - March 2025 | Maturity score - March 2026 |
| --- | --- | --- | --- | --- | --- |
| Engagement of People | 2 | 3 | 0 | 0 | 0 |
| Indentifying Priorities | 1 | 2 | 0 | 0 | 0 |
| Evidence Based Information | 2 | 3 | 0 | 0 | 0 |
| Evidence Synthesis and Guidelines | 2 | 3 | 0 | 0 | 0 |
| Data and Information Systems | 1 | 2 | 0 | 0 | 0 |
| Benchmarking | 1 | 1 | 0 | 0 | 0 |
| Implementation | 1 | 2 | 0 | 0 | 0 |
| Healthcare Improvement | 1 | 2 | 0 | 0 | 0 |

#### Step 2:

Now record which approach you used to evaluate each of the eight components.

Place an x in the appropriate cell to indicate which approach you took (delete and re-enter the data in the table below as appropriate)

- Instinctual - This is a quick rating. Don't worry too much. Just fill in without reference to anything. Later steps will use a more evidence-based approach. This rating method will familiarise you with the process
- Evidence-based with reference to local information/ data - In this approach you use data and information that you have available. This may be surveys, monthly reports, emails. It can be any information that you can refer to that is relevant to your site/ project/ service
- Evidence based with use of evidence-based tools (those provided in the matrix and any others you have used instead) - The tools/ links that are provided are those recommended by our Stakeholders. Choose an area and then refer to the tools. Choose one to start with and see if your approach is consistent with the provided evidence-based approach. Overtime you would plan to look at each of the 8 areas with this approach.

Then indicate how many components were listed in each column.

Add in any notes and or recommendations here.

| **Evidence base** | | | | |
| --- | --- | --- | --- | --- |
| **Domain** | **Component** | **Instinctual** | **Evidence based with reference to local information/ data** | **Evidence based with use of evidence-based tools** |
| **Stakeholder Derived Evidence** | **Engagement of People** | x |  |  |
|  | **Identifying Priorities** | x |  |  |
| **Research Derived Evidence** | **Evidence Based Information** | x |  |  |
|  | **Evidence Synthesis and Guidelines** | x |  |  |
| **Data Derived Evidence** | **Data and Information Systems** | x |  |  |
|  | **Benchmarking** | x |  |  |
| **Implementation Evidence** | **Implementation** | x |  |  |
|  | **Healthcare Improvement** | x |  |  |
| **Count of evidence approaches** |  | 8 | 0 | 0 |

##### Step 3:

##### Evidence based with reference to local information/ data or other evidence-based tools

Where you have accessed some sort of evidence to complete your ranking it is advised to document clearly what they were. Over time it is easy for this information to be lost.

- Document which data sources, tools and resources you used in each of the component areas
- Indicate whether they were sourced from the spreadsheet or whether it was found elsewhere
- Include links to the source - or
- Attach any PDF’s if the data source is likely to change over time
- List information about the data/ information next to the relevant component as listed below:

###### Engagement of People

###### Identifying Priorities

###### Evidence Based Information

###### Evidence Synthesis and Guidelines

###### Data and Information Systems

###### Benchmarking

###### Implementation

###### Healthcare Improvement
