## Supplementary material for "The Monash Learning Health System Maturity Matrix: Codesign of a Tool to Measure and Guide Improvement in Complex Health System Behaviour": S5a_MonashLHSMM_AustralianStrokeLHS_Report_Filled

### Learning Health System Matrix Snapshot report

This document will provide a system/ structure for documentation of use of the matrix over time.

#### Australian Stroke LHS

**Date of evaluation**: 12^th^ November 2024

**Person completing the evaluation**: Darren Rajit

**Role**: Researcher in Residence

#### Step 1:

Complete your matrix rating. (Worksheet 1. Make sure that you alter the cells on row 2, column H, I, J, K, L. This will ensure that the Legend on the spider graph is relevant to your study.)

Then copy and paste your spider graph (From the Summary Sheet) **to replace the one below**.

Australian Stroke LHS

|  | Start of Project |
| --- | --- |
| Engagement of People | 5 |
| Identifying Priorities | 3 |
| Evidence Based Information | 5 |
| Evidence Synthesis and Guidelines | 5 |
| Data and Information Systems | 5 |
| Benchmarking | 5 |
| Implementation | 5 |
| Healthcare Improvement | 4 |

###### Engagement of People

Formation of the Australian Stroke Coalition (ASC) (1) which has been co-led by the consumer advocacy group Stroke Foundation and the Australian and New Zealand Stroke Organisation (formerly Stroke Society of Australasia), representing research and clinician networks since 2008. Codesign as a core principle for knowledge generation is incorporated, with the provision of co-designed modules to educate researchers on how to work with consumers with lived experience, and completion of such modules as a requirement for research grant funding (2). Further, the ASC endorses an “active partnership” model between consumers, clinicians, researchers and broader workforce organisations in all its initiatives e.g. Occupational Therapy Australia (1).

###### Identifying Priorities

Both the ASC and the Australian government have shortlisted priority areas in stroke care quality improvement. National targets for stroke care as proposed by the ASC (55) are informed by priorities areas set within the Acute Stroke Clinical Care Standards (56), which in turn have been developed by a national level committee of interdisciplinary representatives and lived experience representatives (57). However, as there is limited evidence on whether these priorities have been ranked in order to prioritise resources, a ranking beyond “Developing” could not be assigned.

###### Evidence Based Information

Inclusion and generation of research evidence in the Australian Stroke LHS is informed by agreed priority areas within agreed priority populations (58). Additionally, the development of national level standards as in the Acute Stroke Clinical Care Standards has been informed by international and national level clinical guidelines, and systematic reviews and meta-analyses (57). Collectively, this influences downstream grant making efforts (58) and incentivises research generation in priority areas.

###### Evidence Synthesis and Guidelines

Since 2017, the Australian and New Zealand Clinical Guidelines for Stroke Management is a living guideline that continually assesses new published evidence, and updates recommendations following a review and endorsement process (47). Consumers are included in the writing teams for each clinical topic to prioritize recommendations, and also on the Steering Committee (59).

###### Data and Information Systems

The Australian Stroke Data Tool (AuSDaT) (45) was established in 2015 as a single source of integrated stroke data for Australia. The tool enables different data collection programs including the Stroke Foundation Audit program (46) and the Australian Stroke Clinical Registry (AuSCR) which collects patient level clinical care and patient reported outcome measures to be collected (60).

###### Benchmarking

AuSDAT also allows hospitals to collect and monitor data against national level benchmarks. Tailored reports are provided back to participating hospital services (60), whilst the AuSCR allows on-demand access to live interactive dashboards with both peer and national level benchmarks (60)

###### Implementation

There has been demonstrated use of evidence-based implementation strategies and quality improvement methodologies to achieve behaviour change, for example to close evidence-practice gaps (61). Quality improvement programs as interventions have been codesigned and underpinned by theory driven implementation frameworks (62–64). Further, accompanying delivery strategies have been literature informed (20,63). Changes in practice as a result have been routinely captured and reported through existing data infrastructure (AuSCR) (62)

###### Healthcare Improvement

Since 2007, when audit and feedback programs began as part of the Australian Stroke LHS program, there have been significant improvements in service organisation between 1999 and 2019 for access to stroke units, thrombolysis services, and rapid assessment or management for patients with transient ischaemic attack. Analyses of patient-level audits for 2007 to 2019 indicated that the odds of receiving care processes per audit cycle to have significantly increased for thrombolysis, stroke unit access, risk factor advice, and carer training. In parallel, the age-standardised rate of stroke events has reduced by 27% between 2001 and 2020 (65). Further, mortality rates due to stroke has reduced from 37.4 deaths per 100,000 men in 2007 in to 23.8 deaths per 100,000 men in 2021 (67). Likewise, in women, mortality rates have reduced from 36.1 deaths per 100,000 women in 2007 to 23.9 deaths per 100,000 women in 2021 (65). As such there has been marked improvements in stroke related implementation, health system, and patient health outcomes. However, given the relatively recent nature of the implementation of tools and process that are related to LHS principles, such as the transition of the Stroke Guidelines to a living format in 2018 (47), evidence of the ongoing cycles of the LHS are still nascent, with direct implementation process outcomes and program evaluations from such efforts only to be realised in the coming decade. As such a maturity level beyond Established could not be assigned.
