## Supplementary material for "The Monash Learning Health System Maturity Matrix: Codesign of a Tool to Measure and Guide Improvement in Complex Health System Behaviour": S7_MonashLHSMM-NMG-Divergence

Table 1: Summary of high-level changes from the Network Maturity Grid, applied after adaptation and further co-design into the Monash LHS Maturity Matrix (LHS-MM)

|  | **Network Maturity Grid** | **Monash LHS-MM** |
| --- | --- | --- |
| **Underlying LHS Framework** | Constructed from National Academy of Medicine (NAM) description of LHS and additional frameworks (32) | Monash LHS Framework ^7^ |
| **Constructs and Components** | Six Domains with associated components: Systems of Leadership, Governance and Management, Quality Improvement, Community building and Engagement, Data and Analytics, Research | 4 Domains (Stakeholder, Research, Data, Implementation) with associated Components (See Table 3) |
| **Maturity Levels** | 5 Maturity Levels: Not Started, Beginning, Intermediate, Mature, Idealized State | 5 Maturity Levels: Not Started, Beginning, Developing, Established, Transformative |
| **Directions for Usage** | N/A | Can be applied either as a reflexive self-assessment, or as part of a rigorous evaluation |
| **Conceptualisation of the LHS** | As a series of processes across a variety of themes | As system level behaviours across 8 LHS Components, that arise from integrative and localised processes |
| **Incorporation of Theory Driven, Evidence Based Implementation Frameworks** | N/A | Incorporates insights from CFIR ^3^, RE-AIM ^26^ and Implementation Fidelity from Carrol et al. ^27^ |
